# Larazotide Acetate for Treatment of Celiac Disease: A Systematic Review and Meta-Analysis of Randomized Controlled Trials

**DOI:** 10.1101/2020.09.06.20189324

**Authors:** Ahmed Abu-Zaid, Noor Tariq Alhaddab, Razan Abdulkarim Alnujaidi, Hadeel Abdulaziz Alharbi, Fulwah Alangari, Naseem Alyahyawi, Aminah Kamal, Abdulaziz Khalaf Altowairqi, Habeeb Alhabeeb, Sami Almustanyir, Reem Abdullah Alyoubi

## Abstract

**Purpose:** The standard of care for treatment of celiac disease (CD) is a stringent lifetime gluten-free diet (GFD), which is very challenging. Larazotide acetate (AT-1001) is an anti-zonulin which functions as a gut permeability regulator for treatment of CD. We endeavored to conduct a systematic review and meta-analysis of all randomized controlled trials (RCTs) which studied the efficacy and safety of larazotide acetate in patients with CD.

**Methods:** We examined four databases from inception to 20-August-2020 using related keywords. We identified all relevant RCTs and judged their risk of bias. We pooled continuous outcomes as mean difference and dichotomous outcomes as risk ratio with 95% confidence interval under fixed-effects meta-analysis model.

**Results:** Four RCTs met our eligibility criteria, comprising 626 patients (larazotide acetate, n = 465, placebo, n = 161). Three and two studies reported outcomes of patients undergoing gluten challenge and GFD, respectively. For change in lactulose-to-mannitol ratio, the overall effect estimate did not reveal a significant difference between larazotide acetate and placebo groups. For change in total gastrointestinal symptom rating scale (GSRS), subgroup analysis showed that larazotide acetate significantly yielded better symptomatic improvement in the gluten challenge but not gluten free subgroup. Similar finding was found for change in celiac-disease GSRS (CD-GSRS) favoring the gluten challenge over gluten free subgroup. When compared to placebo, larazotide acetate favorably reduced the adverse event (AE) of gluten-related diarrhea in patients who underwent gluten challenge. Other AEs were comparable between both treatment groups.

**Conclusions:** Larazotide acetate is well-endured and superior to placebo in alleviating gastrointestinal symptoms.

## 1. INTRODUCTION

Celiac disease (CD) is a universal public health ailment [1]. The approximated global prevalence of CD is 0.7–1.4% [1]. Biologically, CD is an autoimmune enteropathy prompted by intake of dietary gluten (particularly gliadin peptides) in hereditarily predisposed persons [2]. Despite formerly assumed to involve only the small intestine, CD is presently regarded as a systemic autoimmune disease [3]. CD patients experience a wide array of gastrointestinal and extra-gastrointestinal manifestations [4–6]. Such gastrointestinal manifestations comprise abdominal pain, malabsorption, loss of appetite and long-lasting diarrhea. On the other hand, extra-gastrointestinal manifestations comprise fatigue, anemia, infertility, hepatic disorders, neuropsychiatric symptoms and depression. All in all, CD negatively impacts the quality of welling for CD patients [7] and their caregivers [8].

The standard of care for treatment of CD is a stringent lifetime gluten-free diet (GFD) [3, 9]. Nevertheless, strict compliance with GFD is associated with several challenges. Such challenges include the poor satisfaction of CD patients with their GFD [10]. Interestingly, super adherence to GFD itself has been reported to correlate with poor quality of life for CD patients [11]. Additional challenges include the financial expensiveness and unavailability of gluten-free products when compared to their gluten-containing equivalents [12]. These challenges are further compounded by several reports documenting the intentional and unintentional gluten ingestion by CD patients [7, 13]. Unfortunately, a mere GFD is not adequate to halt the disease pathogenesis or dramatically lessen symptoms in a large proportion of CD patients [14]. Therefore, there is a pressing necessity to devise alternative and novel therapies for CD patients [10].

A key step in the etiopathogenesis of CD involves the mobilization of gliadin peptides from intestinal lumen to lamina propria where gluten-reactive T cells exist, thus triggering a robust adaptive immune response [9]. Two pathways have been proposed for this gliadin peptides trafficking, namely transcellular [15] and paracellular [16] pathways. The transcellular pathway encompasses the co-localization of secretory Immunoglobulin A (IgA) and CD71 to mediate the transcellular transfer of gliadin peptides to lamina propria though transcytosis [15]. On the other hand, the paracellular pathway encompasses the delivery of gliadin peptides to lamina propria through disassembling of tight junctions known as zonula occludens. This disassembling process is mediated by a sequential signaling cascade that involves a key tight junction modulator known as zonulin [16]. In fact, paracellular permeability has been theorized to be an initial promoting event in the etiology of CD [17, 18]. Thus, drugs targeting zonulin hold promising therapeutic potentials for the management of CD.

Larazotide acetate (also recognized as AT-1001) is a novel, synthetic, eight-amino acid peptide that antagonizes zonulin. It is structurally related to the initially isolated zonula occluden toxin (ZOT) generated by *Vibrio cholera* bacterium [9]. Preclinically, larazotide acetate depicted beneficial outcomes in terms of increased tight junction integrity and decreased paracellular permeability in various *in vitro* and *in vivo* experimental models of CD [19–21]. Clinically, larazotide acetate has been scrutinized for tolerability in two phase I clinical trials comprising healthy volunteers [9]. Single and multiple doses of larazotide acetate ranging from as low as 0.25 mg to as high as 36 mg did not result in any severe drug-related side effects and none of the healthy volunteers withdrew from these studies. Nonetheless, mild headache was the most frequently voiced symptom by the healthy volunteers. Overall, these phase I clinical trials in healthy volunteers established the safety of larazotide acetate [9]. Subsequently, the safety and efficacy of larazotide acetate, in various doses, have been examined in four randomized placebo-controlled trials in CD patients in the presence and/or absence of gluten challenge [22–25].

In this study, we attempted to conduct a systematic review and meta-analysis of all randomized placebo-controlled trials that scrutinized the safety and efficacy of larazotide acetate for the management of CD patients.

## 2. METHODS

We carried out this systematic review and meta-analysis in firm adherence to the Preferred Reporting Items for Systematic Reviews and Meta-Analyses (PRISMA) statement [26, 27].

### 2.1. Literature Search

We searched a total of four databases (PubMed, Cochrane Library, Web of Science and Scopus) from inception to August 15th, 2020. We used the following search strategy: (AT1001 OR zot protein, vibrio cholerae OR larazotide acetate) AND (coeliac disease OR gluten enteropath* OR gluten-sensitive enteropath*).

### 2.2. Eligibility Criteria

We selected all publications that met the following criteria for our PICOS evidence-based research question: (I) Patients: individuals with celiac disease undergoing gluten challenge or GFD, (II) Intervention: larazotide acetate, (III) Comparator: placebo, (IV) Outcomes: efficacy and safety endpoints, and (V) Study design: randomized controlled trials. We excluded protocols, animal studies, and abstracts.

### 2.3. Screening of Results

We exported citations using EndNote software to start the screening phase. The screening of results was done through two steps. The first step involved title and abstract screening. The second step involved retrieval of full-text. Additionally, the references of the included studies were examined for potential inclusion of pertinent studies.

### 2.4. Data Extraction

We extracted four major categories of data, namely: (I) baseline demographic and clinical characteristics of the included participants, (II) baseline characteristics of the included studies, (III) efficacy outcomes, and (IV) safety outcomes. Data about baseline demographic and clinical characteristics of included participants comprised sample size, age, female gender, white ethnicity, weight, height, body mass index, time since onset of CD symptoms, time since CD diagnosis and time since start of most recent GFD. Data about baseline characteristics of the included studies comprised http://ClinicalTrials.gov identifier, study design, clinical trial phase, doses, presence of gluten challenge and outcomes assessed in the systematic review and metaanalysis. Data about efficacy outcomes comprised lactulose-to-mannitol (LAMA) ratio [3, 9] and clinical symptoms assessed by total Gastrointestinal Symptom Rating Scale (total GSRS) and CD-specific GSRS (CeD-GSRS) [23–25, 28–30]. Lastly, safety outcomes comprised patients with ≥1 adverse event (AE), ≥1 AE related to study medication, ≥1 severe adverse event, patients who discontinued study medication because of an AE, headache, urinary tract infection, gluten-related flatulence, gluten-related constipation, and gluten-related diarrhea.

### 2.5. Quality Assessment

We used the Cochrane’s risk of bias tool, which is stated in the Cochrane Handbook for Systematic Reviews of Interventions 5.1.0 to examine the risk of bias among the included studies [31]. This tool evaluates the following domains: random sequence generation, allocation concealment, blinding of participants and personnel, blinding of outcome assessment, incomplete outcome data, selective outcome reporting and other potential sources of bias. Two investigators independently assessed the quality of the eligible studies and discrepancies were resolved by a third investigator, if applicable. The investigators’ judgment comprised low, unclear, or high risk of bias for each evaluated domain.

### 2.6. Data Synthesis

Three studies [23–25] used various concentrations of larazotide acetate and we regarded each concentration as a separate study in our meta-analysis. The efficacy endpoints (change in LAMA ratio, total GSRS and CeD-GSRS) were regarded as continuous data, analyzed using the inverse variance method and reported as mean difference (MD) with 95% confidence interval (95% CI) under fixed-effects analysis model. The computation of efficacy endpoints was done using the Review Manager Software version 5.3. The safety outcomes were regarded as dichotomous data, analyzed using the Mantel-Haenszel method and reported as risk ratio (RR) with 95% CI under fixed-effects analysis model. The computation of safety outcomes was done using the Open Meta-Analyst Software. We considered heterogeneity if I-square test (I^2^) > 50% and Chi-Square P< 0.1. When we detected heterogeneity, we performed a sensitivity analysis using the Cochrane’s leave-one-out method.

Regarding individual study data of efficacy endpoints, only the study by Leffler et al. (2012) [23] reported data directly as mean and standard deviation (SD). All other studies reported data as mean and 95% CI and we converted the 95% CI into SD using the methods described in the Cochrane Handbook for systematic reviews of interventions chapter 7.7.3.2. We obtained the 95% CI of studies by extracting the data from their corresponding figures using the WebPlotDigitizer software (http://www.automeris.io/WebPlotDigitizer/), because written values were not provided in the full-texts. The study by Kelly et al. [24] did not provide enough data for calculation of SD; therefore, we estimated the missing SD using the methods described by Weir et al. [32] Furthermore, we performed a subgroup analysis according to the status of gluten intake: gluten challenge (patients receiving gluten diet prior to testing the efficacy of larazotide acetate) and gluten free (patients who remained on a GFD throughout the trial).

## 3. RESULTS

### 3.1 Search results and summary of included studies

Literature search yielded a total of 91 studies after omission of duplicated ones. After title and abstract screening, 83 studies were excluded and the remaining eight studies progressed to full-text screening for eligibility. Finally, a total of four studies met the inclusion criteria and were included in the qualitative and quantitative synthesis [22–25]. **Figure 1** shows the PRIMSA flow diagram. Our meta-analysis included 626 patients; 465 and 161 patients received larazotide acetate and placebo, respectively. Three studies [22–24] reported outcomes of patients undergoing gluten challenge whereas two studies[23, 25] reported outcomes of patients without gluten challenge. The baseline demographic and clinical characteristics of the included participants are depicted in **Table 1**. The baseline characteristics of the included studies are displayed in **Table 2**.

**Figure 1.**
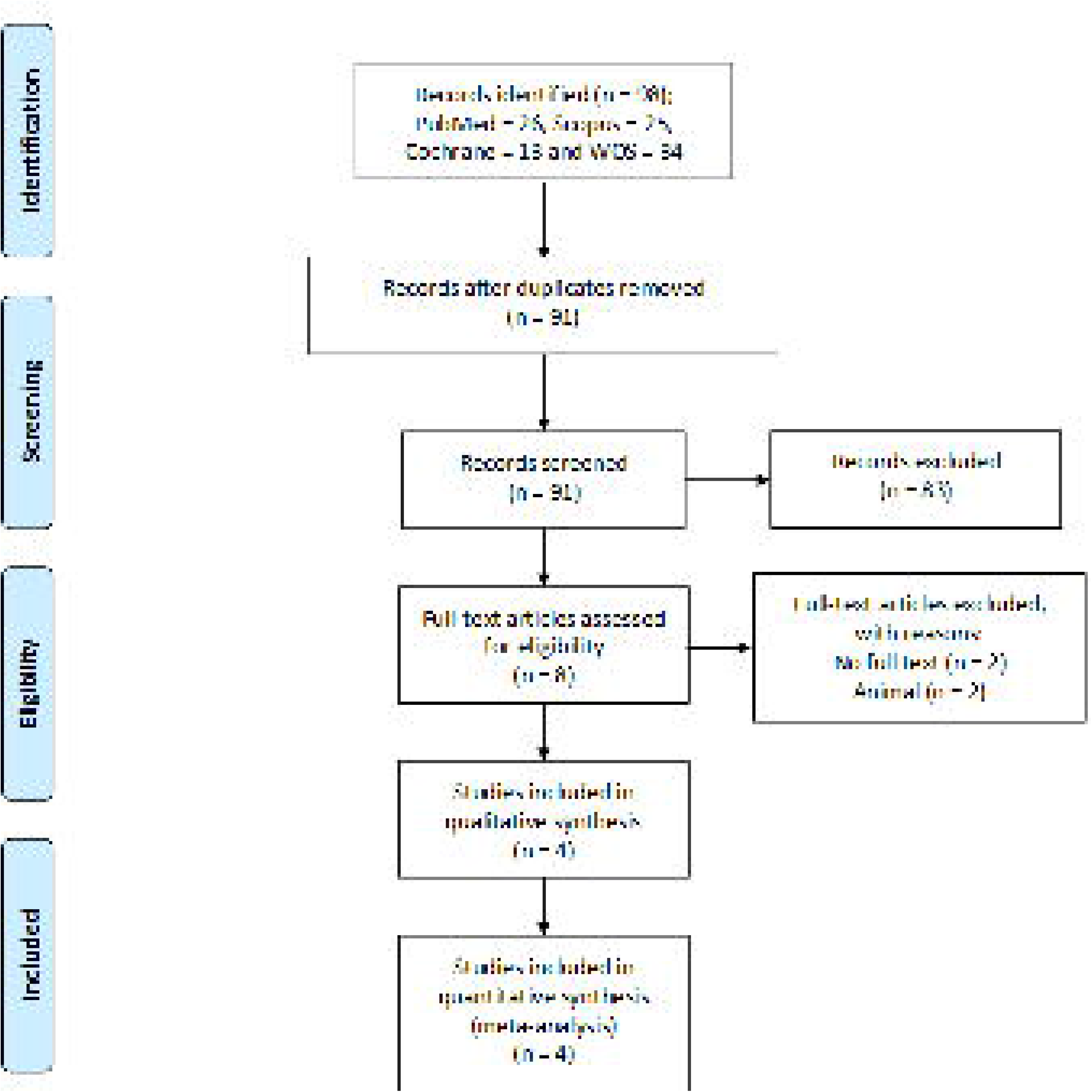
PRISMA flow diagram.

**Table 1.** Baseline demographic and clinical characteristics of the included participants.

| ID | Group | Number | Age, year | Female | White race | Weight, kg | Height, cm | BMI, kg/m2 | Time since CeD symptom onset, month | Time since CeD diagnosis, month | Time on most recent GFD, month |
| --- | --- | --- | --- | --- | --- | --- | --- | --- | --- | --- | --- |
| **Gluten challenge** |  |  |  |  |  |  |  |  |  |  |  |
| Kelly 2013 | 1 mg | 45 | 50.3 [10.09] | 36 (80%) | 45 (100%) | 70.5 [12.72] | 166.8 [9.51] | 25.3 [3.80] | 139.8 [144.61] | 66.3 [52.53] | 68.4 [58.80] |
|  | 4 mg | 44 | 49.5 [12.42] | 31 (70%) | 44 (100%) | 75.5 [17.07] | 168.1 [8.84] | 26.7 [5.43] | 155.0 [190.10] | 60.0 [65.54] | 81.0 [116.21] |
|  | 8 mg | 44 | 47.4 [13.61] | 31 (70%) | 44 (100%) | 73.3 [13.83] | 169.1 [7.60] | 25.6 [4.16] | 132.1 [138.25] | 54.0 [43.52] | 63.3 [53.62] |
|  | Placebo | 44 | 50.3 [10.24] | 29 (66%) | 44 (100%) | 74.0 [15.25] | 170.9 [9.20] | 25.2 [4.02] | 143.2 [162.47] | 63.4 [52.08] | 68.7 [57.44] |
| Leffler 2012 | 0.25 mg | 12 | 46.3 | 31 (53%) | - | - | - | - | - | - | - |
|  | 1 mg | 12 |  |  | - | - | - | - | - | - | - |
|  | 4 mg | 12 |  |  | - | - | - | - | - | - | - |
|  | 8 mg | 12 |  |  | - | - | - | - | - | - | - |
|  | Placebo | 14 |  |  | - | - | - | - | - | - | - |
| Paterson 2007 | 12 mg | 14 | 37.6 (8.1) | 12 (85.7) | - | 61.9 (12) | 167 [7.6] | 22 [3.3] | - | - | - |
|  | Placebo | 7 | 33.3 [9.9] | 5 (71.4) | - | 71.3 [12.6] | 169 [7.2] | 25 [3.5] | - | - | - |
| **Gluten-free diet** |  |  |  |  |  |  |  |  |  |  |  |
| Leffler 2015 | 0.5 mg | 86 | 44.2 [14.4] | 73 (84.9%) | 84 (97.7%) | 75.5 [16.0] | 165.4 [8.9] | 27.6 [5.3] | 149.2 [144.8] | 58.0 [50.1] | 60.3 [60.1] |
|  | 1.0 mg | 85 | 46.2 [15.0] | 71 (83.5%) | 85 (100%) | 72.2 [15.4] | 166.2 [8.6] | 26.0 [4.6] | 162.9 [150.6] | 66.7 [68.2] | 71.4 [86.2] |
|  | 2.0 mg | 87 | 44.7 [16.0] | 72 (82.8%) | 86 (98.9%) | 73.4 [15.3] | 167.4 [8.5] | 26.1 [4.6] | 174.6 [174.7] | 71.5 [65.07] | 70.9 [63.7] |
|  | Placebo | 84 | 45.5 [14.6] | 69 (82.1%) | 83 (98.8%) | 74.3 [16.8] | 167.3 [9.6] | 26.6 [5.4] | 145.5 [141.1] | 60.3 [51.3] | 62.1 [58.7] |
| Leffler 2012 | 8 mg | 12 | 46.3 | 45(53%) | - | - | - | - | - | - | - |
|  | Placebo | 12 |  |  | - | - | - | - | - | - | - |

Data of female and white race are expressed as number (percentage), otherwise data are expressed as mean [standard deviation].
BMI: body mass index; CeD: celiac disease; GFD: gluten-free diet

**Table 2.** Baseline characteristics of the included studies.

| Study | ClinicalTrials.gov | Phase | Doses | Gluten challenge | Outcomes assessed in systemic review and meta-analysis |
| --- | --- | --- | --- | --- | --- |
| Paterson 2007 | NCT00362856 | II | 12 mg | Yes | Change in LAMA ratio, patients with $\geq 1$ adverse event, patients with $\geq 1$ adverse event related to study medication, patients with $\geq 1$ severe adverse event, patients who discontinued study medication because of an adverse event, gluten-related diarrhea, gluten-related flatulence, gluten-related constipation |
| Kelly 2013 | NCT004929601 | II | 1, 4, 8 mg | Yes | Change in LAMA ratio, change in total GSRS score, change in total CeD-GSRS score, patients with $\geq 1$ adverse event, patients with $\geq 1$ adverse event related to study medication, patients with $\geq 1$ severe adverse event, patients who discontinued study medication because of an adverse event, headache, gluten-related diarrhea, gluten-related flatulence and gluten-related constipation |
| Leffler 2012 | NCT00362856 | II | 0.25, 1, 4, 8 mg | Yes<br>One cohort [8 mg] did not | Change in LAMA ratio, change in total GSRS score, change in total CeD-GSRS score, patients with $\geq 1$ adverse event, patients with $\geq 1$ adverse event related to study medication, patients with $\geq 1$ severe adverse event, patients who discontinued study medication because of an adverse event, headache. |
| Leffler 2015 | NCT01396213 | II | 0.5, 1, 2 mg | No | Change in total GSRS score, change in total CeD-GSRS score, Patients with $\geq 1$ adverse event, headache, urinary tract infection. |

CeD-GSRS: celiac disease gastrointestinal rating symptoms scale; GSRS: gastrointestinal rating symptoms scale; ID: identifier; LAMA: lactulose-to-mannitol ratio;
NCT: national clinical trial identifier as per ClinicalTrials.gov

### 3.2. Quality assessment of the included studies

Overall, the included studies were of moderate-to-high quality and had low risk of bias. The method of randomization was not mentioned in the study by Leffler et al. 2012 [23]; thus, we judged the ‘random generation sequence’ domain as unclear risk. Additionally, we judged the ‘blinding outcome assessment’ domain as high risk in the study by Kelly et al. [33]. All studies were supported by sponsors, therefore, we judged ‘other bias’ as high risk for trial sponsorship. The graph and summary of risk of bias are illustrated in **Figure 2**.

**Figure 2.**
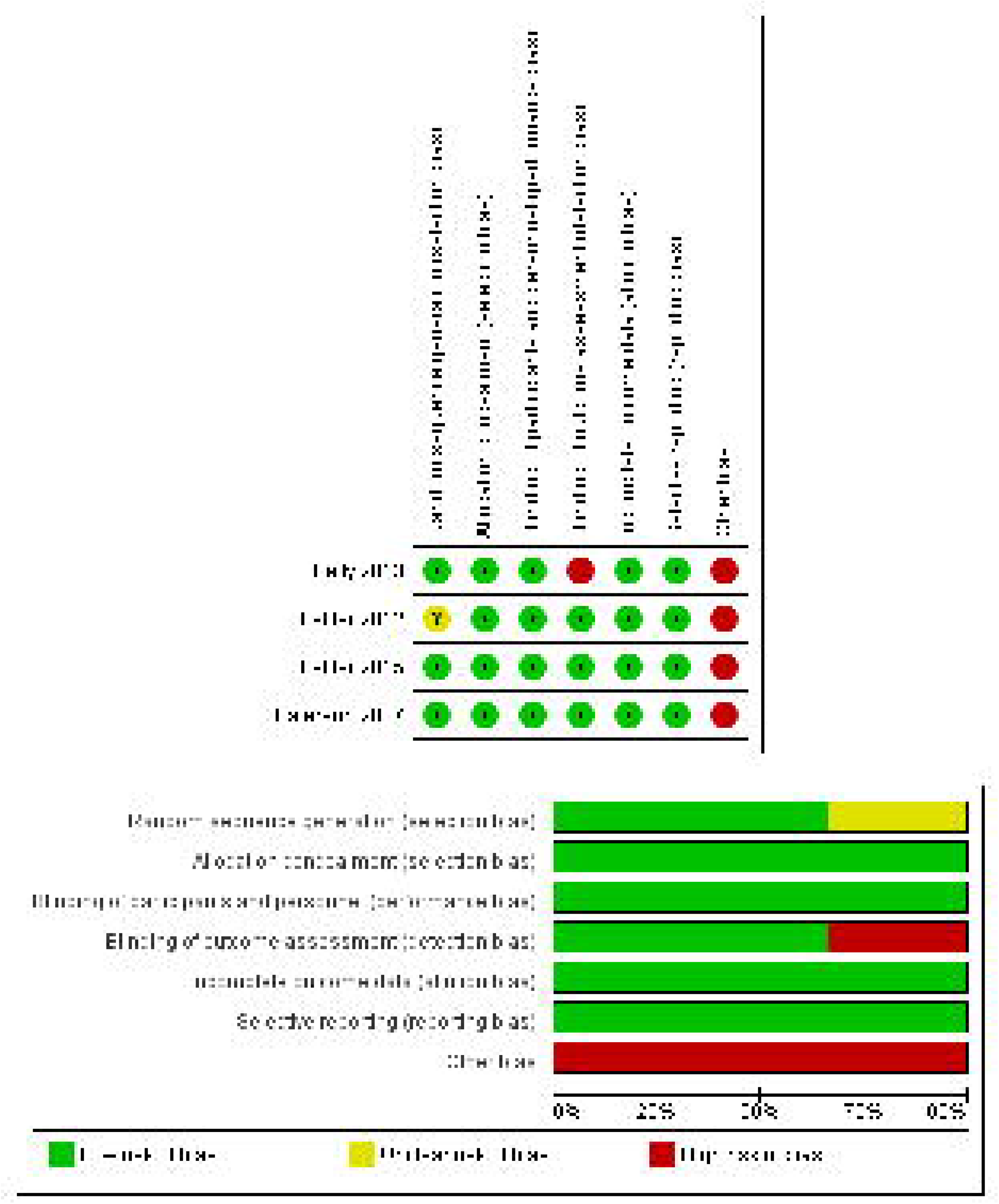
Risk of bias summary and graph.

### 3.3. Analysis of efficacy outcomes

#### 3.3.1 Change in LAMA ratio

Three studies were meta-analyzed [22–24]. The overall effect estimate did not reveal a significant difference between larazotide acetate and placebo groups (MD = –0.11, 95% CI [-0.43, 0.20], p = 0.48). Pooled analysis was homogeneous (l^2^ = 0%, P = 1). Additionally, subgroup analysis revealed no significant difference between larazotide acetate and placebo groups for the gluten challenge (MD = –0.17, 95% CI [-0.51, 0.18], p = 0.35; I^2^ = 0%, P = 1.00) and gluten-free (MD = 0.15, 95% CI [-0.64, 0.95], p = 0.7; heterogeneity: not applicable) subgroups **(Figure 3)**.

**Figure 3.**
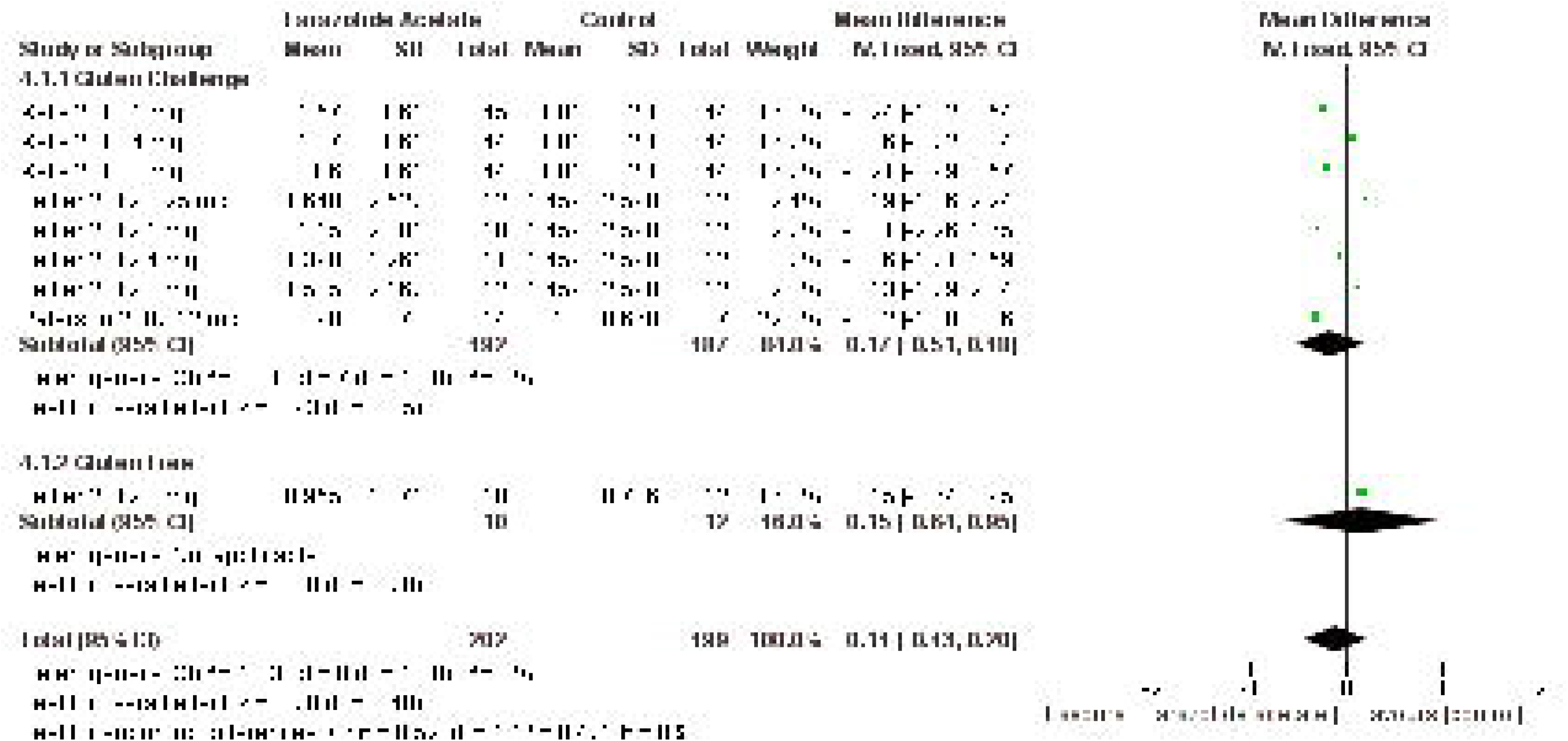
Forest plot showing the change in lactulose-to-mannitol (LAMA) ratio in among disease patients with gluten challenge.

#### 3.3.2. Change in total GSRS score

Three studies were meta-analyzed [23–25]. The overall effect estimate did not indicate a substantial difference between larazotide acetate and placebo groups (MD = –0.09, 95% Cl [0.22, 0.04], p = 0.19). Pooled analysis was homogeneous (l^2^ = 0%, P = 0.55). However, subgroup analysis showed that larazotide acetate significantly yielded better symptomatic improvement in the gluten challenge (MD = –0.20, 95% CI [-0.40, –0.01], p = 0.04) but not gluten free (MD = 0.01, 95% CI [-0.17, 0.19], p = 0.84) subgroup. Pooled analysis was homogenous (l^2^ = 0%, P = 0.46 and I^2^ = 0%, P = 0.90, respectively) **(Figure 4)**.

**Figure 4.**
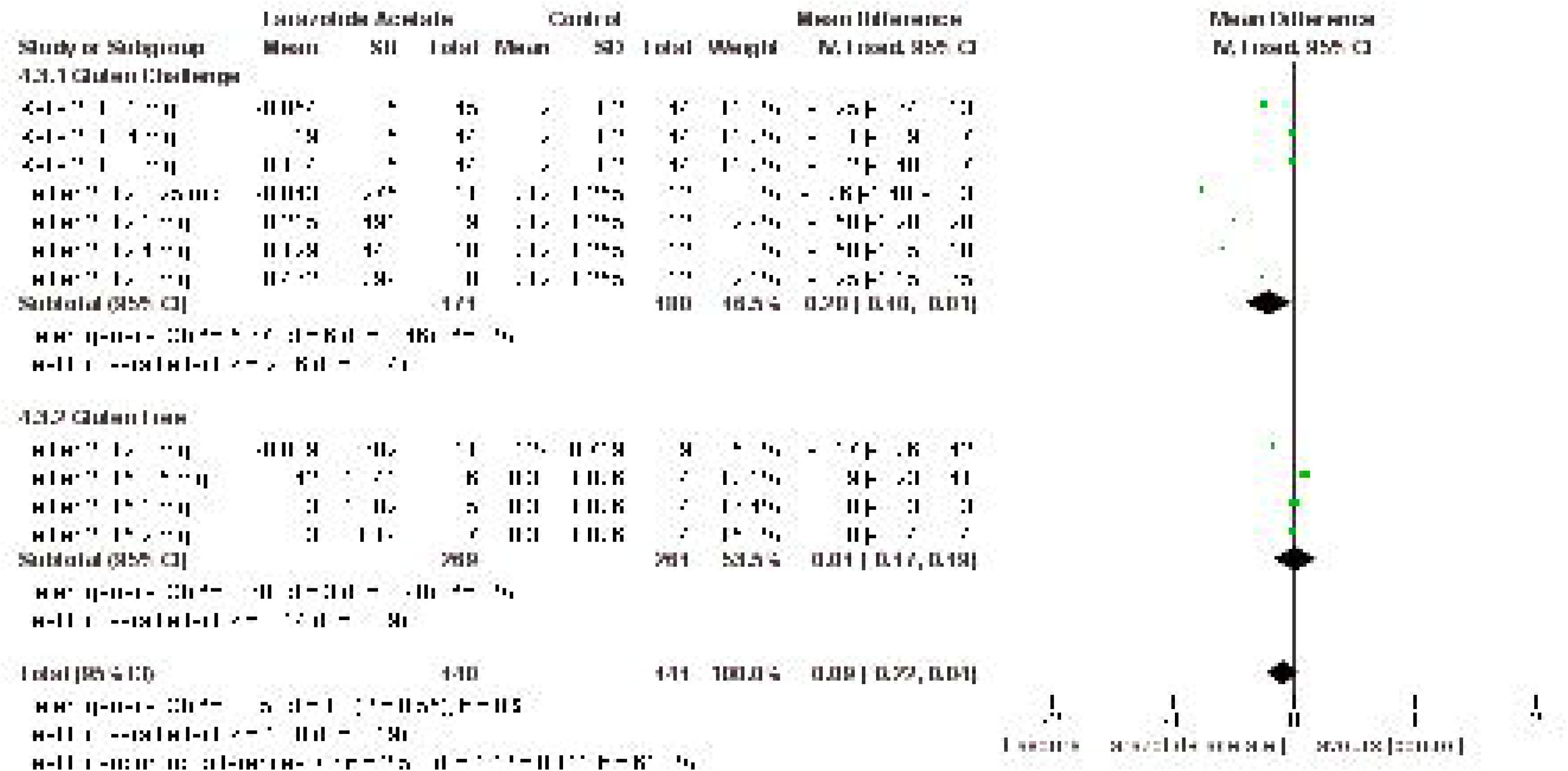
Forest plot showing the change in total Gastrointestinal Symptom Rating Scale (GSRS) among celiac disease patients.

#### 3.3.3. Change in total CeD-GSRS score

Three studies were meta-analyzed [23–25]. The overall effect estimate did not demonstrate a major difference between larazotide acetate and placebo groups (MD = –0.06, 95% CI [-0.20, 0.07], p = 0.36). Pooled analysis was homogeneous (l^2^ = 20%, P = 0.25). However, subgroup analysis demonstrated that larazotide acetate substantially offered superior symptomatic improvement in the gluten challenge (MD = –0.26, 95% CI [-0.49, –0.03], p = 0.03) but the not gluten free (MD = 0.04, 95% CI [-0.12, 0.21], p = 0.6) subgroup. Pooled analysis was homogenous (l^2^ = 9%, P = 0.36 and I^2^ = 0%, P = 0.68, respectively) **(Figure 5)**.

**Figure 5.**
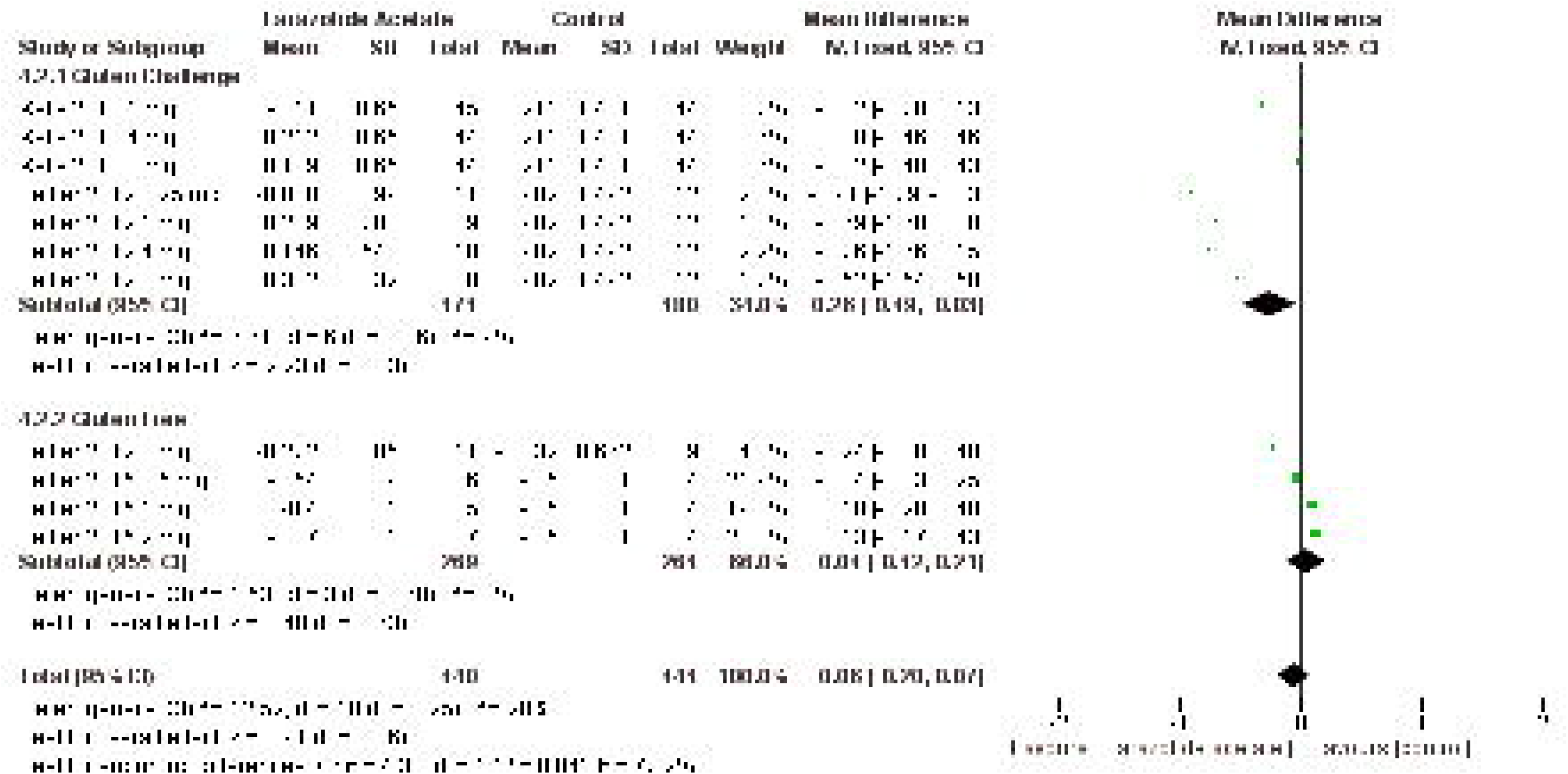
Forest plot showing the change in total Celiac Disease-Gastrointestinal Symptom Rating Scale (CeD-GSRS) among celiac disease patients.

### 3.4. Analysis of Safety Endpoints

#### 3.4.1. Gluten challenge subgroup

For patients who underwent gluten challenge, three studies were meta-analyzed for safety outcomes [23–25]. When compared to placebo, larazotide acetate favorably reduced the AE of gluten-related diarrhea in patients who underwent gluten challenge (RR = 0.420, 95% CI [0.246, 0.717], p = 0.001) **(Figure 6)**. Pooled analysis was homogenous (P = 0.211).

**Figure 6.**
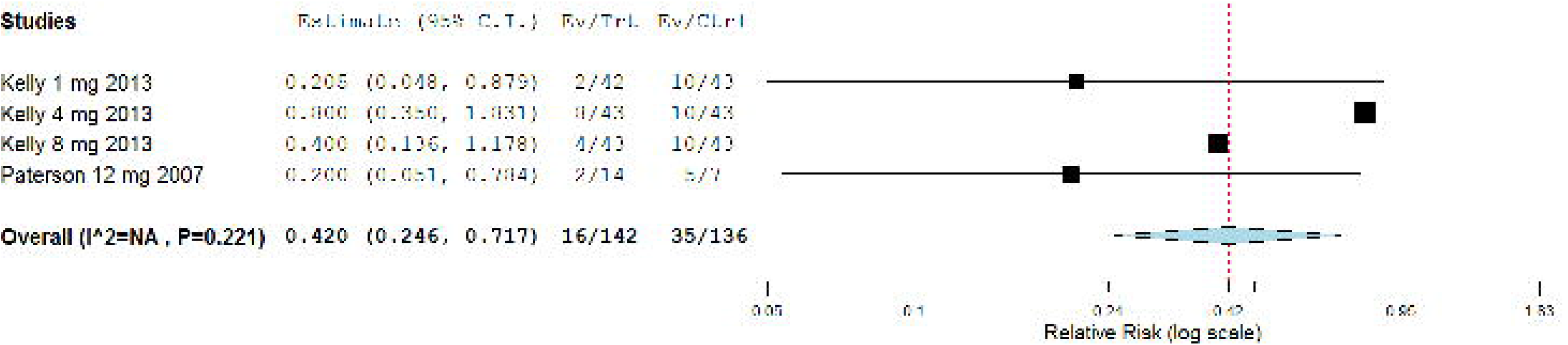
Forest plot showing the safety outcome of gluten-related diarrhea among celiac disease patients with gluten challenge.

On the other hand, the overall RR presented no significant difference between larazotide acetate and placebo groups regarding patients with ≥1 AE (RR = 0.953, 95% CI [0.844, 1.076], p = 0.435) **(Figure 7)**, patients with ≥1 AE related to study medication (RR = 0.853, 95% CI [0.664, 1.896], p = 0.214) **(Figure 8)**, patients with ≥1 severe AE (RR = 0.815, 95% CI [0.365, 1.821], p = 0.618) **(Figure 9)**, patients who discontinued study medication because of an AE (RR = 0.754, 95% CI [0.390, 1.460], p = 0.403) **(Figure 10)**, headache (RR = 1.045, 95% CI [0.538, 2.030], p = 0.897) **(Figure 11)**, gluten-related flatulence (RR = 0.703, 95% Cl [0.0.356, 1.388], p = 0.310) **(Figure 12)** and gluten-related constipation (RR = 0.778, 95% CI [0.322, 1.883], p = 0.578) **(Figure 13)**. Pooled analysis was homogenous (P>0.1) under fixed-effects model, except for patients with ≥ 1 severe AE, which was heterogeneous (P = 0.094); thus results were analyzed under the random-effects model.

**Figure 7.**
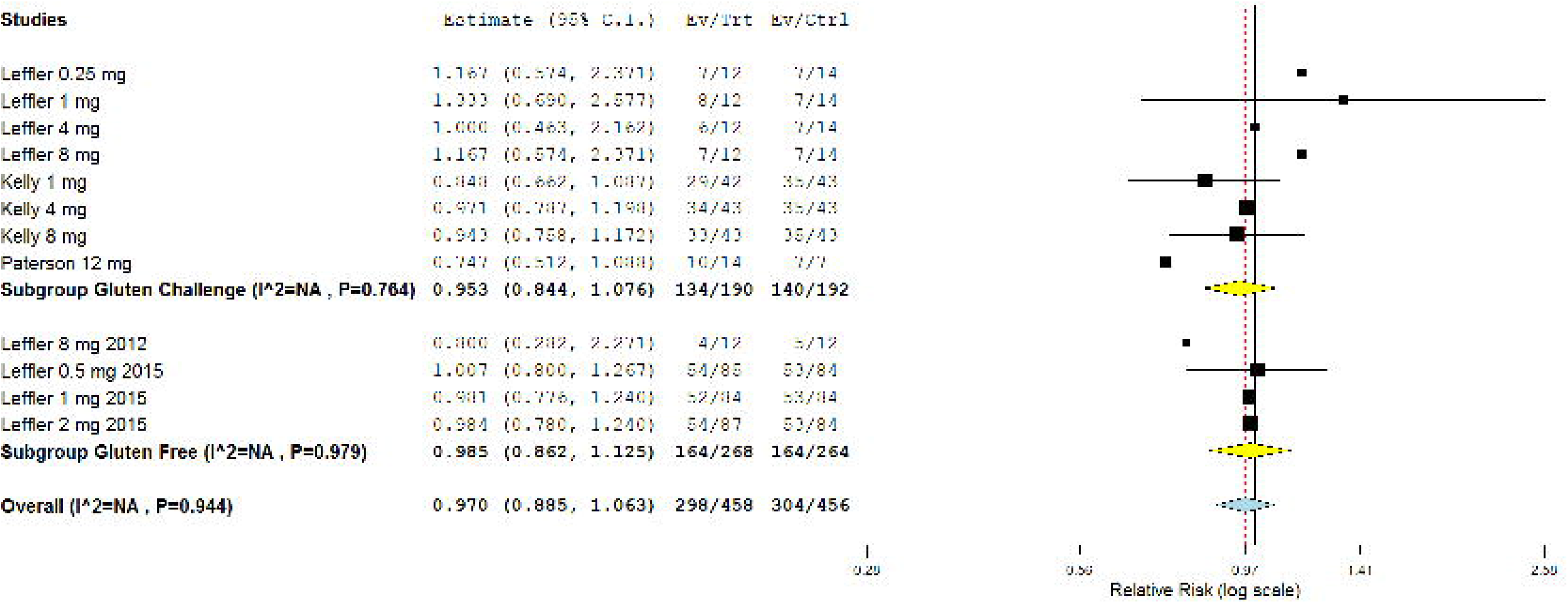
Forest plot showing the safety outcome of ≥ 1 adverse event among celiac disease patients.

**Figure 8.**
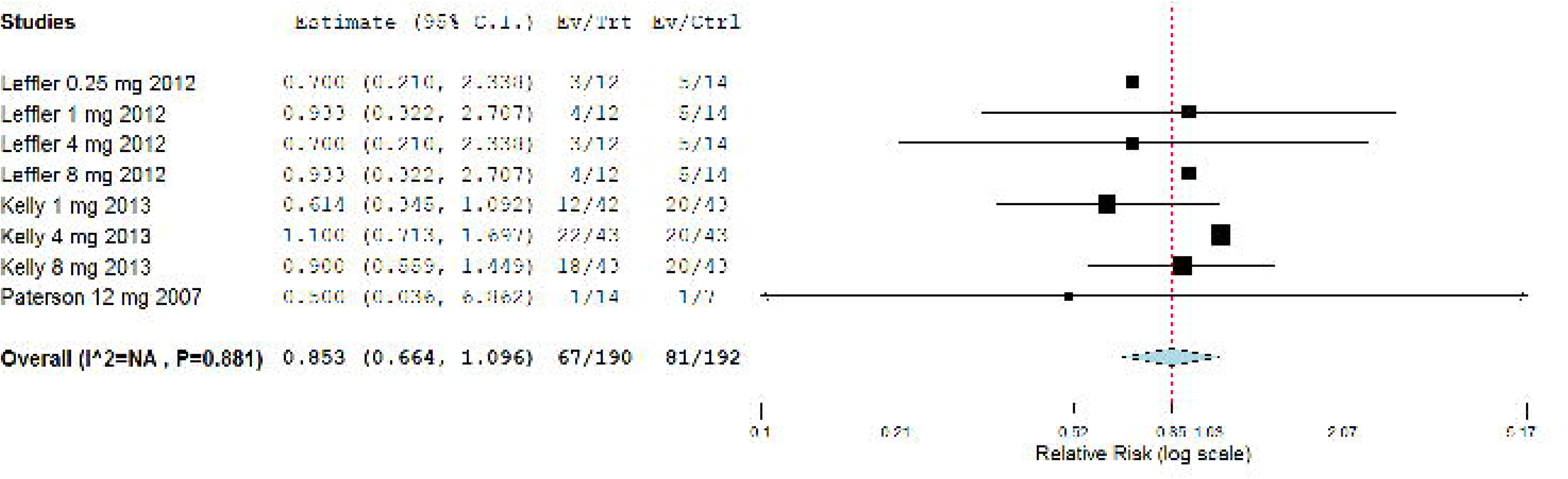
Forest plot showing the safety outcome of ≥ 1 adverse event related to study treatment (deemed so by the investigator) among celiac disease patients with gluten challenge.

**Figure 9.**
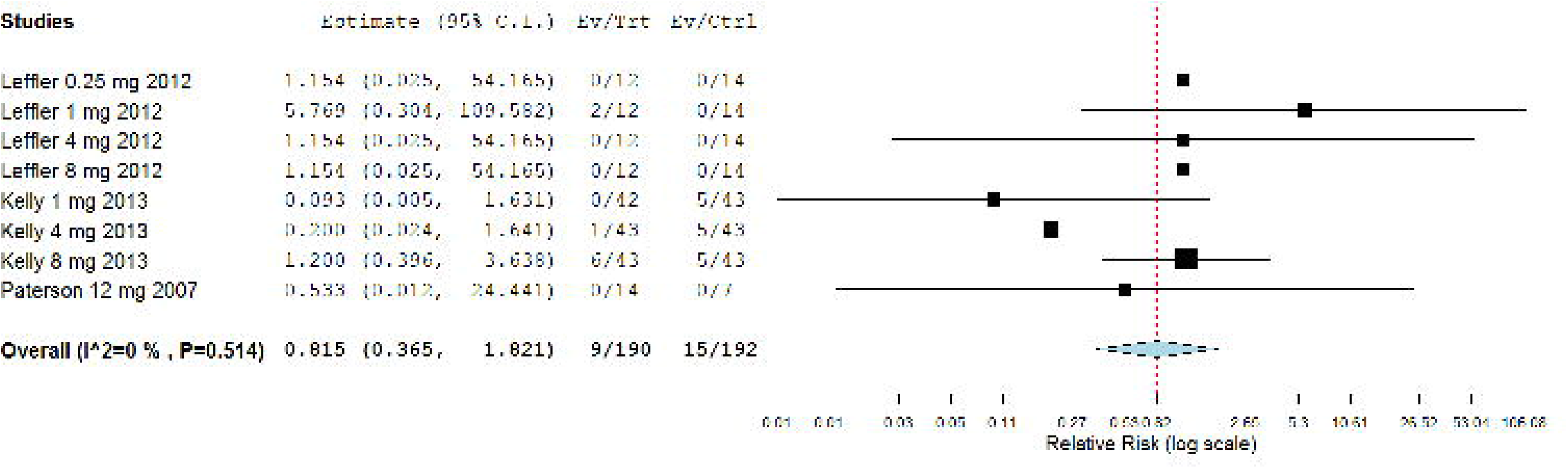
Forest plot showing the safety outcome of ≥ 1 severe adverse event among celiac disease patients with gluten challenge.

**Figure 10.**
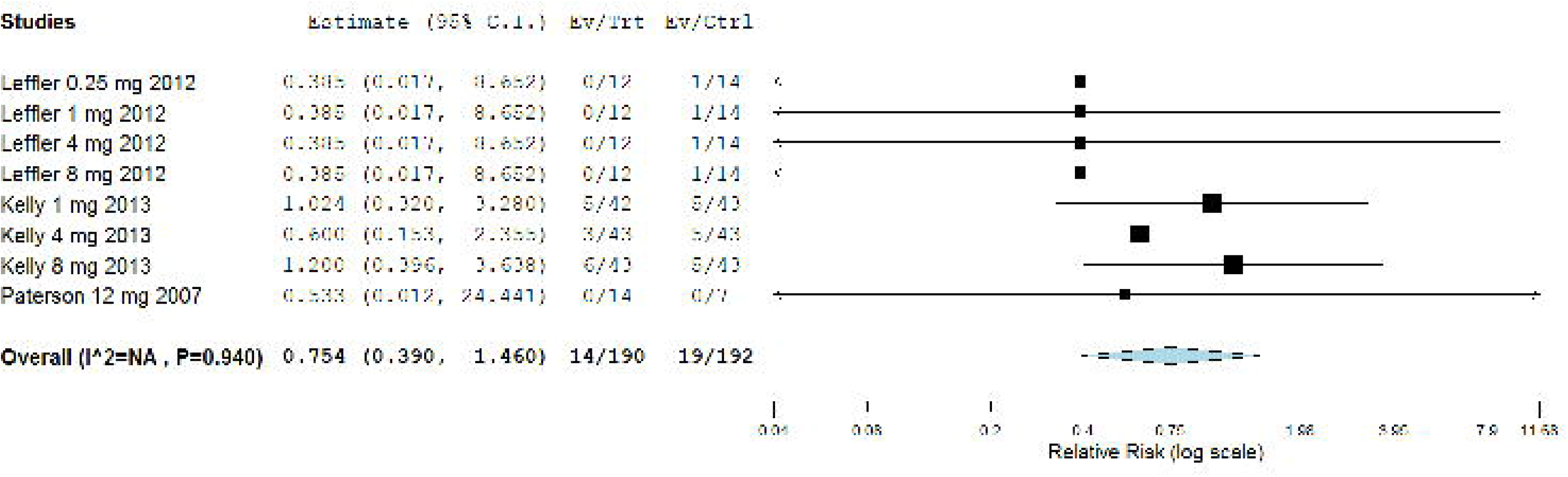
Forest plot showing the safety outcome of stopping treatment due to an adverse event among celiac disease patients with gluten challenge.

**Figure 11.**
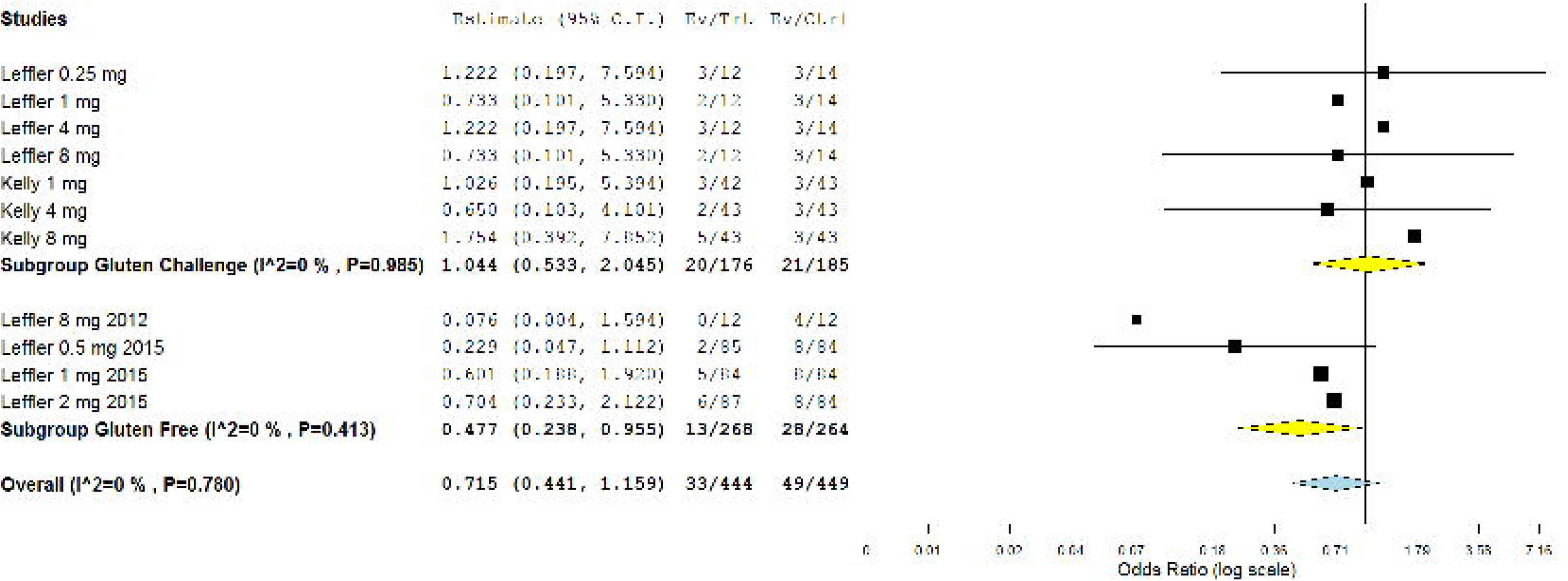
Forest plot showing the safety outcome of headache among celiac disease patients.

**Figure 12.**
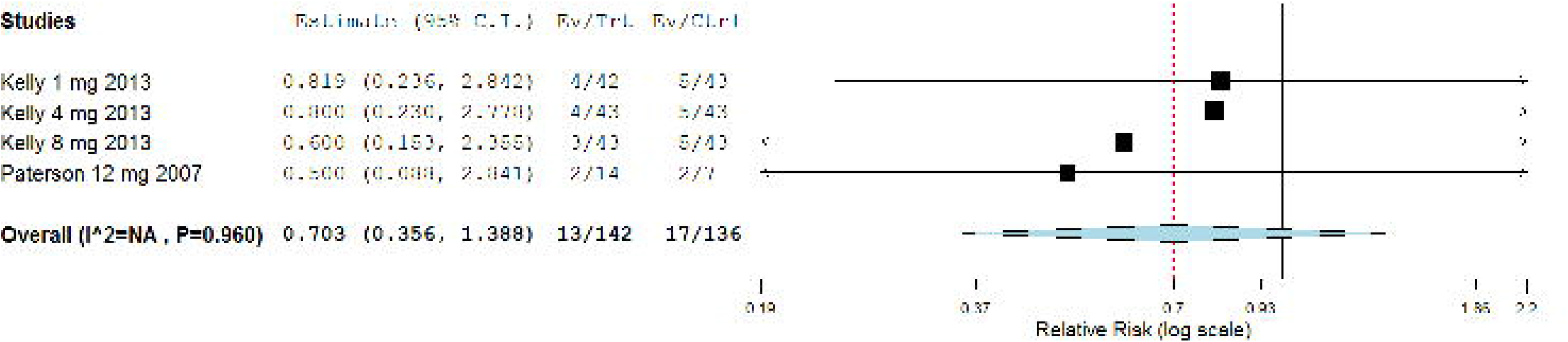
Forest plot showing the safety outcome of gluten-related flatulence among celiac disease patients with gluten challenge.

**Figure 13.**
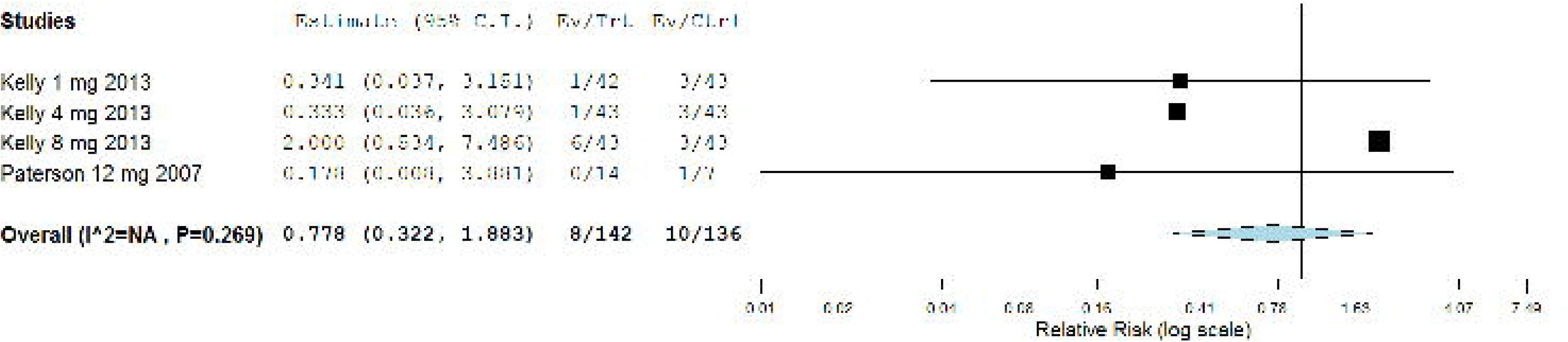
Forest plot showing the safety outcome of gluten-related constipation among celiac disease patients with gluten challenge.

#### 3.4.2. Gluten-free subgroup

For gluten free patients who did not undergo gluten challenge, two studies were meta-analyzed for safety outcomes [23, 25]. When compared to placebo, larazotide acetate favorably reduced the AE of headache (RR = 0.432, 95% CI [0.220, 0.851], p = 0.015) **(Figure 11)**. Pooled analysis was homogenous (P = 0.401). Conversely, the overall RR revealed no significant variance between larazotide acetate and placebo groups with regard to patients with ≥1 AE (RR = 0.985, 95% CI [0.862, 1.125], p = 0.822) **(Figure 7)** and urinary tract infection (RR = 1.142, 95% CI [0.388, 3.662], p = 0.809) **(Figure 14)**. Pooled analysis was homogeneous (P = 0.979 and P = 0.414, respectively).

**Figure 14.**
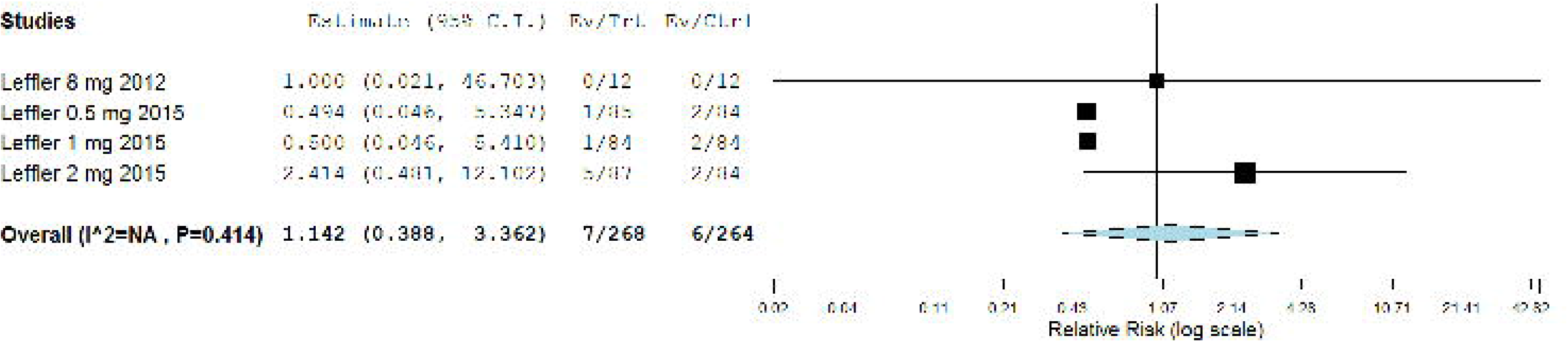
Forest plot showing the safety outcome of urinary tract infection among celiac disease patients without gluten challenge.

## 4. DISCUSSION

### 4.1. Summary of findings

With regard to larazotide acetate versus placebo for management of CD patients, we included four studies comprising a total of 626 patients (465 and 161 patients received larazotide acetate and placebo, respectively). Results obtained from this study are clinically important in consideration of the moderate-to-high quality of the included randomized controlled trials.

For CD patients undergoing gluten challenge, pooled analysis of the change in urinary LAMA ratio failed to demonstrate superiority of larazotide acetate over placebo in reducing intestinal permeability. The lack of this efficacy outcome could be attributed to the high inconsistency in LAMA ratio measurements which were obtained at somewhat uncontrolled outpatient settings. While LAMA ratio is the most frequently utilized investigation to gauge the functional permeability of the gut [3, 9]. Nonetheless, it is bound by several technical limitations that ought to be acknowledged [34–36]. For example, sampling period and concomitant intake of liquid/food can adversely interfere with the assayed levels of urinary sugars, namely lactulose and mannitol.

The impaired intestinal mucosal barrier in CD contributes to the gut leakiness–inflammation circlet that gives rise to an array of gastrointestinal and extra-gastrointestinal symptoms [37]. Total GSRS and CeD-GSRS are validated metrics used commonly in clinical settings to evaluate CD patients with gastrointestinal symptoms [23–25, 28–30]. In CD patients undergoing gluten challenge, pooled analysis of total GSRS and CeD-GSRS substantially favored larazotide acetate over placebo in subsiding symptoms. Interestingly, improvements in both scales were not substantially attained in CD patients undergoing GFD. The lack of larazotide acetate efficacy in producing considerable symptomatic improvement among CD patients undergoing GFD cannot be explained and this matter warrants further investigation.

CD adversely impacts the quality of life of its patients [7]. Interestingly, despite the symptomatic improvement among CD patients undergoing gluten challenge and receiving larazotide acetate, two studies reported no significant difference in quality of life [23, 24]. Quality of life was assessed using the validated Psychological General Well-Being Index (PGWBI) which has been used previously in patients with CD [28, 38].

The vast majority of studies used different concentrations of larazotide acetate ranging from as low as 0.25 mg to as high as 12 mg. Intriguingly, close inspection of the efficacy outcomes depicted an inverse dose-outcome correlation. The lower doses were more effective when compared to the higher ones. This remark is not unique to larazotide acetate and is actually noticed in other orally ingested peptide drugs, such as linaclotide. Linaclotide is a medication utilized for the management of patients with irritable bowel syndrome with predominate constipation [39, 40]. Maximal efficacy of linaclotide was more observed with lower doses when contrasted to higher ones. Proposed elucidations for these antipodal dose-outcome effects comprise drug accumulation and receptor desensitization at higher doses [9].

Larazotide acetate exhibited a well-endured safety profile when compared to placebo. Diarrhea is a major distressing symptom in patient with CD [4–6]. In patients undergoing gluten challenge, gluten-related diarrhea was substantially reduced in patients who received larazotide acetate. Notably, there was no clear relationship between drug dose and side effects. The optimal dose of larazotide acetate is yet to be determined.

### 4.2. Clinical implications, scientific perspectives and future directions

At present time, there are no United States Food and Drug Administration (FDA)-greenlighted drugs for management of CD [9]. GFD is the standard of care for the treatment of CD [3, 9]. Its strict adherence by patients is very challenging [10] and does not amply pause disease pathogenesis or reduce symptoms [14]. Thus, there is an unmet requisite for non-dietary therapies for treatment of patients with CD.

Larazotide acetate surfaces as a novel alternative therapy for treatment of CD. Our study shows that larazotide acetate is well-tolerated and improves symptoms in patients with CD. Mechanistically, larazotide acetate lessens the delivery of immunogenic gliadin peptides to the mucosal gluten-reactive T cells. While larazotide acetate blocks the transport of gliadin peptides to lamina propria through paracellular pathway, it does not affect the transcellular pathway [9]. It does not completely reestablish tolerance or fully inhibit immune responses to gluten. Intake of larazotide acetate does not protect against intentional ingestion of gluten-containing products. Akin to other luminal active drugs, it is a short-acting drug and thus will mandate repetitive dosing before patients ingest gluten. Overall, larazotide acetate is less probable to offer a definitive cure to patients with CD. Thus, larazotide acetate is likely to be proposed as a long-term maintenance therapy and it should be administered to complement rather than displace the standard of care GFD. With regard to safety profile, larazotide acetate is largely safer when compared to other immunotherapeutic drugs which interfere with immune responses and could result in undesirable off-target effects.

In line with the consistent symptomatic improvement induced by larazotide acetate, there is an ongoing large-sized, randomized, double-blinded, placebo-controlled phase III trial that will examine the tolerability and efficacy of larazotide acetate in relieving CD symptoms (http://ClinicalTrials.gov Identifier: NCT03569007). Indeed, larazotide acetate is one of the very few drugs that entered phase III trials and on its way for potential FDA approval.

### 4.3. Strengths and limitations

Our study holds several strengths. Most importantly, this is the first systematic review and meta-analysis report that pooled the efficacy and safety outcomes of larazotide acetate for the management of patients with CD. Whenever applicable, we performed subgroup analysis according to presence or absence of gluten challenge. Nevertheless, our study is not free of limitations. Such limitations include the relative small number of included studies and their respective small sample size. While the data are encouraging with respect to symptomatic amelioration, important data on histologic and autoantibody (for example, anti-endomysial and anti-tissue transglutaminase) improvements are lacking and these are major curtails. Additionally, there were heterogeneities in drug doses and efficacy endpoints which could have negatively affected the overall results. This matter of heterogeneity in outcomes has been voiced recently in a publication that called for meticulous assessment and reporting of endpoints to enhance transparency and comparability of therapeutic trials pertaining to CD [41]. This matter is particularly true for measurement of change in LAMA ratio among other efficacy endpoints.

## 5. Conclusions

This systematic review and meta-analysis advocates that in patients with CD, larazotide acetate is superior to placebo in alleviating gastrointestinal symptoms with a well-endured safety profile. Larazotide acetate may be coadministered to complement rather than displace the standard of care GFD. Lastly, large-sized studies with longer follow up and appropriate efficacy endpoint measurements—such as urinary LAMA ratio, autoantibodies and histology, are needed to solidify the efficacy and tolerability of larazotide acetate in patients with CD. The ongoing phase III trial will provide very meaningful findings once its results are communicated.

## Data Availability

The data used to support the findings of this study are included within the article.

## COMPLIANCE WITH ETHICAL STANDARDS

## Funding

None.

## Conflict of interest

None.

## Ethics approval

Not required for this study type.

## Consent to participate

Not applicable.

## Consent for publication

Not applicable.

## Code availability

Not applicable.

## Authors’ contributions

AA contributed to study supervision, design, conception, analysis and manuscript writing. NTA, RAA, HAA, FA, NA, AK, AKA, HA, SA and RAA contributed to literature review and data collection. All authors edited manuscript for editorial and intellectual contents. All authors read and approved the final draft of manuscript for submission.

## Notes

### Competing Interest Statement

The authors have declared no competing interest.

### Author Declarations

Not applicable or required.

